## Supplementary material for "Comparative genomic study for revealing the complete scenario of COVID-19 pandemic in Bangladesh": Supplementary File 2.pdf

We gratefully acknowledge the following Authors from the Originating laboratories responsible for obtaining the specimens, as well as the Submitting laboratories where the genome data were generated and shared via GISAID, on which this research is based.

All Submitters of data may be contacted directly via www.gisaid.org

Authors are sorted alphabetically.

| Accession ID | Originating Laboratory | Submitting Laboratory | Authors |
|---|---|---|---|
| EPI_ISL_1360425, EPI_ISL_1360426, EPI_ISL_1360427, EPI_ISL_1360428, EPI_ISL_1360430, EPI_ISL_1360446, EPI_ISL_1360447, EPI_ISL_1360448, EPI_ISL_1360449, EPI_ISL_1360450, EPI_ISL_1360451, EPI_ISL_1469964, EPI_ISL_1469965, EPI_ISL_1469966, EPI_ISL_1469967, EPI_ISL_1469968, EPI_ISL_1469969, EPI_ISL_1469970, EPI_ISL_1469971, EPI_ISL_1469972, EPI_ISL_1469973, EPI_ISL_1469974, EPI_ISL_1469975, EPI_ISL_1469976, EPI_ISL_1469977, EPI_ISL_1469978, EPI_ISL_1469979, EPI_ISL_1469986 | | | |
| see above | Child Health Research Foundation | Child Health Research Foundation | CHRF Bangladesh Genomics Team |
| EPI_ISL_1498126, EPI_ISL_1498127, EPI_ISL_1498131, EPI_ISL_1498132, EPI_ISL_1498136, EPI_ISL_1498137, EPI_ISL_1498138, EPI_ISL_1498139, EPI_ISL_1498140, EPI_ISL_1498141, EPI_ISL_1498145, EPI_ISL_1498149, EPI_ISL_1498150, EPI_ISL_1498151 | | | |
| see above | Institute for Developing Science and Health Initiatives (ideSHi) | Institute for Developing Science and Health Initiatives (ideSHi) | Hassan Afrad, Sadia Rahman, Fidausi Qadri, Tahmina Shirin |
| EPI_ISL_1508828, EPI_ISL_1508833, EPI_ISL_1508841 | National Institute of Laboratory Medicine and Referral Center | Genomic Research Lab, BCSIR | Md. Murshed Hasan Sarkar, Shahina Akter, Abu Sayeed Mohammad Mahmud, Mohammad Samir Uzzaman, Eshrar Osman, Md. Ahasan Habib, Tanjina Akhter Banu, Barna Goswami, Iffat Jahan, Md. Saddam Hossain, Tasnim Nafisa, Md. Maruf Ahmed Molla, Mahmuda Yeasmin, Asish Kumar Ghosh, Arifa Akram, A. K. M. Shamsuzzaman, Md. Salim Khan |
| EPI_ISL_1508891, EPI_ISL_1508892, EPI_ISL_1508893, EPI_ISL_1508915 | National Institute of Laboratory Medicine and Referral Center | Genomic Research Lab. BCSIR | Md. Murshed Hasan Sarkar, Shahina Akter, Abu Sayeed Mohammad Mahmud, Mohammad Samir Uzzaman, Eshrar Osman, Md. Ahasan Habib, Tanjina Akhter Banu, Barna Goswami, Iffat Jahan, Md. Saddam Hossain, Tasnim Nafisa, Md. Maruf Ahmed Molla, Mahmuda Yeasmin, Asish Kumar Ghosh, Arifa Akram, A. K. M. Shamsuzzaman, Md. Salim Khan |
| EPI_ISL_1508940, EPI_ISL_1508942, EPI_ISL_1508943, EPI_ISL_1508946, EPI_ISL_1508950, EPI_ISL_1508952, EPI_ISL_1508954, EPI_ISL_1508955, EPI_ISL_1508999 | National Institute of Laboratory Medicine and Referral Center | Genomic Research Lab, BCSIR | Md. Murshed Hasan Sarkar, Shahina Akter, Abu Sayeed Mohammad Mahmud, Mohammad Samir Uzzaman, Eshrar Osman, Md. Ahasan Habib, Tanjina Akhter Banu, Barna Goswami, Iffat Jahan, Md. Saddam Hossain, Tasnim Nafisa, Md. Maruf Ahmed Molla, Mahmuda Yeasmin, Asish Kumar Ghosh, Arifa Akram, A. K. M. Shamsuzzaman, Md. Salim Khan |
| EPI_ISL_1509000, EPI_ISL_1509001, EPI_ISL_1509272 | National Institute of Laboratory Medicine and Referral Center | Genomic Research Lab, BCSIR | Tasnim Nafisa, Md. Murshed Hasan Sarkar, Shahina Akter, Abu Sayeed Mohammad Mahmud, Mohammad Samir Uzzaman, Eshrar Osman, Md. Ahasan Habib, Tanjina Akhter Banu, Barna Goswami, Iffat Jahan, Md. Saddam Hossain, Md. Maruf Ahmed Molla, Mahmuda Yeasmin, Asish Kumar Ghosh, Arifa Akram, A. K. M. Shamsuzzaman, Md. Salim Khan |
| EPI_ISL_1509298, EPI_ISL_1509299, EPI_ISL_1509637, EPI_ISL_1509924, EPI_ISL_1520103, EPI_ISL_1520105 | National Institute of Laboratory Medicine and Referral Center | Genomic Research Lab, BCSIR | Md. Maruf Ahmed Molla, Md. Murshed Hasan Sarkar, Shahina Akter, Abu Sayeed Mohammad Mahmud, Mohammad Samir Uzzaman, Eshrar Osman, Md. Ahasan Habib, Tanjina Akhter Banu, Barna Goswami, Iffat Jahan, Md. Saddam Hossain, Tasnim Nafisa, Mahmuda Yeasmin, Asish Kumar Ghosh, Arifa Akram, A. K. M. Shamsuzzaman, Md. Salim Khan |
| EPI_ISL_1524771, EPI_ISL_1524775, EPI_ISL_1524776, EPI_ISL_1524802, EPI_ISL_1524872 | National Institute of Laboratory Medicine and Referral Center | Genomic Research Lab, BCSIR | Md. Murshed Hasan Sarkar, Shahina Akter, Abu Sayeed Mohammad Mahmud, Mohammad Samir Uzzaman, Eshrar Osman, Md. Ahasan Habib, Tanjina Akhter Banu, Barna Goswami, Iffat Jahan, Md. Saddam Hossain, Tasnim Nafisa, Md. Maruf Ahmed Molla, Mahmuda Yeasmin, Asish Kumar Ghosh, Arifa Akram, A. K. M. Shamsuzzaman, Md. Salim Khan |
| EPI_ISL_1531533, EPI_ISL_1531534, EPI_ISL_1531553, EPI_ISL_1531554, EPI_ISL_1531555 | National Institute of Laboratory Medicine and Referral Center | Genomic Research Lab, BCSIR | Md. Murshed Hasan Sarkar, Shahina Akter, Abu Sayeed Mohammad Mahmud, Mohammad Samir Uzzaman, Eshrar Osman, Md. Ahasan Habib, Tanjina Akhter Banu, Barna Goswami, Iffat Jahan, Md. Saddam Hossain, Tasnim Nafisa, Md. Maruf Ahmed Molla, Mahmuda Yeasmin, Asish Kumar Ghosh, Arifa Akram, A. K. M. Shamsuzzaman, Md. Salim Khan |
| EPI_ISL_1531556, EPI_ISL_1531557, EPI_ISL_1531558 | National Institute of Laboratory Medicine and Referral Center | Genomic Research Lab, BCSIR | Md. Murshed Hasan Sarkar, Shahina Akter, Abu Sayeed Mohammad Mahmud, Mohammad Samir Uzzaman, Eshrar Osman, Md. Ahasan Habib, Tanjina Akhter Banu, Barna Goswami, Iffat Jahan, Md. Saddam Hossain, Tasnim Nafisa, Md. Maruf Ahmed Molla, Mahmuda Yeasmin, Asish Kumar Ghosh, Arifa Akram, A. K. M. Shamsuzzaman, Md. Salim Khan |
| EPI_ISL_1531559, EPI_ISL_1531560 | National Institute of Laboratory Medicine and Referral Center | Genomic Research Lab, BCSIR | Shahina Akter, Md. Murshed Hasan Sarkar, Abu Sayeed Mohammad Mahmud, Mohammad Samir Uzzaman, Eshrar Osman, Md. Ahasan Habib, Tanjina Akhter Banu, Barna Goswami, Iffat Jahan, Md. Saddam Hossain, Tasnim Nafisa, Md. Maruf Ahmed Molla, Mahmuda Yeasmin, Asish Kumar Ghosh, Arifa Akram, A. K. M. Shamsuzzaman, Md. Salim Khan |
| EPI_ISL_1531561, EPI_ISL_1531562 | National Institute of Laboratory Medicine and Referral Center | Genomic Research Lab, BCSIR | Tanjina Akhter Banu, Md. Murshed Hasan Sarkar, Abu Sayeed Mohammad Mahmud, Mohammad Samir Uzzaman, Eshrar Osman, Md. Ahasan Habib, Shahina Akter, Barna Goswami, Iffat Jahan, Md. Saddam Hossain, Mohammad Mohi Uddin, Tasnim Nafisa, Md. Maruf Ahmed Molla, Mahmuda Yeasmin, Asish Kumar Ghosh, Arifa Akram, A. K. M. Shamsuzzaman, Md. Salim Khan |
| EPI_ISL_1531563, EPI_ISL_1531564 | National Institute of Laboratory Medicine and Referral Center | Genomic Research Lab, BCSIR | Barna Goswami, Md. Murshed Hasan Sarkar, Abu Sayeed Mohammad Mahmud, Mohammad Samir Uzzaman, Eshrar Osman, Md. Ahasan Habib, Shahina Akter, Tanjina Akhter Banu, Iffat Jahan, Md. Saddam Hossain, Mohammad Mohi Uddin, Tasnim Nafisa, Md. Maruf Ahmed Molla, Mahmuda Yeasmin, Asish Kumar Ghosh, Arifa Akram, A. K. M. Shamsuzzaman, Md. Salim Khan |
| EPI_ISL_1531565, EPI_ISL_1531566, EPI_ISL_1531567 | National Institute of Laboratory Medicine and Referral Center | Genomic Research Lab, BCSIR | Iffat Jahan, Md. Murshed Hasan Sarkar, Abu Sayeed Mohammad Mahmud, Mohammad Samir Uzzaman, Eshrar Osman, Md. Ahasan Habib, Shahina Akter, Tanjina Akhter Banu, Barna Goswami, Md. Saddam Hossain, Mohammad Mohi Uddin, Tasnim Nafisa, Md. Maruf Ahmed Molla, Mahmuda Yeasmin, Asish Kumar Ghosh, Arifa Akram, A. K. M. Shamsuzzaman, Md. Salim Khan |
| EPI_ISL_1531568, EPI_ISL_1531569 | National Institute of Laboratory Medicine and Referral Center | Genomic Research Lab, BCSIR | Md. Saddam Hossain, Md. Murshed Hasan Sarkar, Abu Sayeed Mohammad Mahmud, Mohammad Samir Uzzaman, Eshrar Osman, Md. Ahasan Habib, Shahina Akter, Tanjina Akhter Banu, Barna Goswami, Iffat Jahan, Mohammad Mohi Uddin, Tasnim Nafisa, Md. Maruf Ahmed Molla, Mahmuda Yeasmin, Asish Kumar Ghosh, Arifa Akram, A. K. M. Shamsuzzaman, Md. Salim Khan |
| EPI_ISL_1531570, EPI_ISL_1531571, EPI_ISL_1531572, EPI_ISL_1531735 | National Institute of Laboratory Medicine and Referral Center | Genomic Research Lab, BCSIR | Md. Saddam Hossain, Md. Murshed Hasan Sarkar, Abu Sayeed Mohammad Mahmud, Mohammad Samir Uzzaman, Eshrar Osman, Md. Ahasan Habib, Shahina Akter, Tanjina Akhter Banu, Barna Goswami, Iffat Jahan, Md. Saddam Hossain, Mohammad Mohi Uddin, Tasnim Nafisa, Md. Maruf Ahmed Molla, Mahmuda Yeasmin, Asish Kumar Ghosh, Arifa Akram, A. K. M. Shamsuzzaman, Md. Salim Khan |
| EPI_ISL_1531743, EPI_ISL_1531744, EPI_ISL_1531776, EPI_ISL_1532065 | National Institute of Laboratory Medicine and Referral Center | Genomic Research Lab, BCSIR | Md. Murshed Hasan Sarkar, Abu Sayeed Mohammad Mahmud, Mohammad Samir Uzzaman, Eshrar Osman, Md. Ahasan Habib, Shahina Akter, Tanjina Akhter Banu, Barna Goswami, Iffat Jahan, Md. Saddam Hossain, Mohammad Mohi Uddin, Tasnim Nafisa, Md. Maruf Ahmed Molla, Mahmuda Yeasmin, Asish Kumar Ghosh, Arifa Akram, A. K. M. Shamsuzzaman, Md. Salim Khan |
| EPI_ISL_1532855, EPI_ISL_1532982, EPI_ISL_1533423, EPI_ISL_1533426, EPI_ISL_1533427 | National Institute of Laboratory Medicine and Referral Center | Genomic Research Lab, BCSIR | Md. Murshed Hasan Sarkar, Shahina Akter, Abu Sayeed Mohammad Mahmud, Mohammad Samir Uzzaman, Eshrar Osman, Md. Ahasan Habib, Tanjina Akhter Banu, Barna Goswami, Iffat Jahan, Md. Saddam Hossain, Mohammad Mohi Uddin, Tasnim Nafisa, Md. Maruf Ahmed Molla, Mahmuda Yeasmin, Asish Kumar Ghosh, Arifa Akram, A. K. M. Shamsuzzaman, Md. Salim Khan |
| EPI_ISL_1533429, EPI_ISL_1533430, EPI_ISL_1533728 | National Institute of Laboratory Medicine and Referral Center | Genomic Research Lab, BCSIR | Abu Sayeed Mohammad Mahmud, Md. Murshed Hasan Sarkar, Mohammad Samir Uzzaman, Eshrar Osman, Md. Ahasan Habib, Shahina Akter, Tanjina Akhter Banu, Barna Goswami, Iffat Jahan, Md. Saddam Hossain, Mohammad Mohi Uddin, Tasnim Nafisa, Md. Maruf Ahmed Molla, Mahmuda Yeasmin, Asish Kumar Ghosh, Arifa Akram, A. K. M. Shamsuzzaman, Md. Salim Khan |
| EPI_ISL_1533802, EPI_ISL_1533852 | National Institute of Laboratory Medicine and Referral Center | Genomic Research Lab, BCSIR | Mohammad Mohi Uddin, Md. Murshed Hasan Sarkar, Mohammad Samir Uzzaman, Eshrar Osman, Md. Ahasan Habib, Shahina Akter, Tanjina Akhter Banu, Abu Sayeed Mohammad Mahmud, Barna Goswami, Iffat Jahan, Md. Saddam Hossain, Mohammad Mohi Uddin, Tasnim Nafisa, Md. Maruf Ahmed Molla, Mahmuda Yeasmin, Asish Kumar Ghosh, Arifa Akram, A. K. M. Shamsuzzaman, Md. Salim Khan |
| EPI_ISL_1534610 | National Institute of Laboratory Medicine and Referral Center | Genomic Research Lab, BCSIR | Md. Murshed Hasan Sarkar, Shahina Akter, Abu Sayeed Mohammad Mahmud, Mohammad Samir Uzzaman, Eshrar Osman, Md. Ahasan Habib, Tanjina Akhter Banu, Barna Goswami, Iffat Jahan, Md. Saddam Hossain, Mohammad Mohi Uddin, Tasnim Nafisa, Md. Maruf Ahmed Molla, Mahmuda Yeasmin, Asish Kumar Ghosh, Arifa Akram, A. K. M. Shamsuzzaman, Md. Salim Khan |

| | | | |
|---|---|---|---|
| EPI_ISL_1534611 | National Institute of Laboratory Medicine and Referral Center | Genomic Research Lab, BCSIR | Uzzaman, Eshrar Osman, Md. Ahasan Habib, Tanjina Akhter Banu, Barna Goswami, Iffat Jahan, Md. Saddam Hossain, Mohammad Mohi Uddin, Tasnim Nafisa, Md. Maruf Ahmed Molla, Mahmuda Yeasmin, Asish Kumar Ghosh, Arifa Akram, A. K. M. Shamsuzzaman, Md. Salim Khan |
| EPI_ISL_1534612, EPI_ISL_1534613 | National Institute of Laboratory Medicine and Referral Center | Genomic Research Lab, BCSIR | Md. Murshed Hasan Sarkar, Shahina Akter, Abu Sayeed Mohammad Mahmud, Mohammad Samir Uzzaman, Eshrar Osman, Md. Ahasan Habib, Tanjina Akhter Banu, Barna Goswami, Iffat Jahan, Md. Saddam Hossain, Mohammad Mohi Uddin, Tasnim Nafisa, Md. Maruf Ahmed Molla, Mahmuda Yeasmin, Asish Kumar Ghosh, Arifa Akram, A. K. M. Shamsuzzaman, Md. Salim Khan |
| EPI_ISL_1534614, EPI_ISL_1534615, EPI_ISL_1534616 | National Institute of Laboratory Medicine and Referral Center | Genomic Research Lab, BCSIR | Md. Maruf Ahmed Molla, Md. Murshed Hasan Sarkar, Shahina Akter, Abu Sayeed Mohammad Mahmud, Mohammad Samir Uzzaman, Eshrar Osman, Md. Ahasan Habib, Tanjina Akhter Banu, Barna Goswami, Iffat Jahan, Md. Saddam Hossain, Mohammad Mohi Uddin, Tasnim Nafisa, Mahmuda Yeasmin, Asish Kumar Ghosh, Arifa Akram, A. K. M. Shamsuzzaman, Md. Salim Khan |
| EPI_ISL_1534617, EPI_ISL_1534618, EPI_ISL_1534619 | National Institute of Laboratory Medicine and Referral Center | Genomic Research Lab, BCSIR | Tasnim Nafisa, Md. Murshed Hasan Sarkar, Shahina Akter, Abu Sayeed Mohammad Mahmud, Mohammad Samir Uzzaman, Eshrar Osman, Md. Ahasan Habib, Tanjina Akhter Banu, Barna Goswami, Iffat Jahan, Md. Saddam Hossain, Mohammad Mohi Uddin, Md. Maruf Ahmed Molla, Mahmuda Yeasmin, Asish Kumar Ghosh, Arifa Akram, A. K. M. Shamsuzzaman, Md. Salim Khan |
| EPI_ISL_1534620, EPI_ISL_1534621 | National Institute of Laboratory Medicine and Referral Center | Genomic Research Lab, BCSIR | Shahina Akter, Md. Murshed Hasan Sarkar, Abu Sayeed Mohammad Mahmud, Mohammad Samir Uzzaman, Eshrar Osman, Md. Ahasan Habib, Tanjina Akhter Banu, Barna Goswami, Iffat Jahan, Md. Saddam Hossain, Mohammad Mohi Uddin, Tasnim Nafisa, Md. Maruf Ahmed Molla, Mahmuda Yeasmin, Asish Kumar Ghosh, Arifa Akram, A. K. M. Shamsuzzaman, Md. Salim Khan |
| EPI_ISL_1538405 | National Institute of Laboratory Medicine and Referral Center | Dr. Qudrat-I-Khuda Road, Dhaka-1205, Bangladesh | Tanjina Akhter Banu, Md. Murshed Hasan Sarkar, Abu Sayeed Mohammad Mahmud, Mohammad Samir Uzzaman, Eshrar Osman, Md. Ahasan Habib, Shahina Akter, Barna Goswami, Iffat Jahan, Md. Saddam Hossain, Mohammad Mohi Uddin, Tasnim Nafisa, Md. Maruf Ahmed Molla, Mahmuda Yeasmin, Asish Kumar Ghosh, Arifa Akram, A. K. M. Shamsuzzaman, Md. Salim Khan |
| EPI_ISL_1538406 | National Institute of Laboratory Medicine and Referral Center | Genomic Research Lab, BCSIR | Tanjina Akhter Banu, Md. Murshed Hasan Sarkar, Abu Sayeed Mohammad Mahmud, Mohammad Samir Uzzaman, Eshrar Osman, Md. Ahasan Habib, Shahina Akter, Barna Goswami, Iffat Jahan, Md. Saddam Hossain, Mohammad Mohi Uddin, Tasnim Nafisa, Md. Maruf Ahmed Molla, Mahmuda Yeasmin, Asish Kumar Ghosh, Arifa Akram, A. K. M. Shamsuzzaman, Md. Salim Khan |
| EPI_ISL_1538407, EPI_ISL_1538408, EPI_ISL_1538409, EPI_ISL_1538410, EPI_ISL_1538411 | National Institute of Laboratory Medicine and Referral Center | Genomic Research Lab, BCSIR | Md. Murshed Hasan Sarkar, Abu Sayeed Mohammad Mahmud, Mohammad Samir Uzzaman, Eshrar Osman, Md. Ahasan Habib, Shahina Akter, Tanjina Akhter Banu, Barna Goswami, Iffat Jahan, Md. Saddam Hossain, Mohammad Mohi Uddin, Tasnim Nafisa, Md. Maruf Ahmed Molla, Mahmuda Yeasmin, Asish Kumar Ghosh, Arifa Akram, A. K. M. Shamsuzzaman, Md. Salim Khan |
| EPI_ISL_1547365 | Department of Genetic Engineering and Biotechnology, Shahjalal University of Science and Technology | Genomic Research Lab, BCSIR | Iffat Jahan, Md. Murshed Hasan Sarkar, Abu Sayeed Mohammad Mahmud, Mohammad Samir Uzzaman, Eshrar Osman, Md. Ahasan Habib, Shahina Akter, Tanjina Akhter Banu, Barna Goswami, Md. Saddam Hossain, Mohammad Mohi Uddin, Md. Shamsul Haque Prodhan, Md. Hammadul Hoque, G. M. Nurnabi Azad Jewel, Md. Nazmul Hasan, Md. Fahmid Hossain Bhuiyan, Md. Asraful Jahan, Ajit Ghosh, Md. Akkas Ali, Md. Salim Khan |
| EPI_ISL_1547372 | Department of Genetic Engineering and Biotechnology, Shahjalal University of Science and Technology | Genomic Research Lab, BCSIR | Mohammad Mohi Uddin, Md. Murshed Hasan Sarkar, Abu Sayeed Mohammad Mahmud, Mohammad Samir Uzzaman, Eshrar Osman, Md. Ahasan Habib, Shahina Akter, Tanjina Akhter Banu, Barna Goswami, Iffat Jahan, Md. Saddam Hossain, Md. Kamrul Islam, Md. Shamsul Haque Prodhan, Md. Hammadul Hoque, G. M. Nurnabi Azad Jewel, Md. Nazmul Hasan, Md. Fahmid Hossain Bhuiyan, Md. Asraful Jahan, Ajit Ghosh, Md. Akkas Ali, Md. Salim Khan |
| EPI_ISL_1547828, EPI_ISL_1548044, EPI_ISL_1548059, EPI_ISL_1548072 | Department of Genetic Engineering and Biotechnology, Shahjalal University of Science and Technology | Genomic Research Lab, BCSIR | Md. Murshed Hasan Sarkar, Abu Sayeed Mohammad Mahmud, Mohammad Samir Uzzaman, Eshrar Osman, Md. Ahasan Habib, Shahina Akter, Tanjina Akhter Banu, Barna Goswami, Iffat Jahan, Md. Saddam Hossain, Mohammad Mohi Uddin, Md. Kamrul Islam, Md. Shamsul Haque Prodhan, Md. Hammadul Hoque, G. M. Nurnabi Azad Jewel, Md. Nazmul Hasan, Md. Fahmid Hossain Bhuiyan, Md. Asraful Jahan, Ajit Ghosh, Md. Akkas Ali, Md. Salim Khan |
| EPI_ISL_1575127 | Institute for Developing Science and Health Initiatives (ideSHi) | Institute for Developing Science and Health Initiatives (ideSHi) | Hassan Afrad, Sadia Rahman, Fidausi Qadri, Tahmina Shirin |
| EPI_ISL_1582390, EPI_ISL_1582391, EPI_ISL_1582392, EPI_ISL_1582393, EPI_ISL_1582394, EPI_ISL_1582395, EPI_ISL_1582396, EPI_ISL_1582397 | Institute of Epidemiology, Disease Control and Research (IEDCR) | Institute for Developing Science and Health Initiatives (ideSHi) | Hassan Afrad, Sadia Rahman, Fidausi Qadri, Tahmina Shirin |
| EPI_ISL_1583158, EPI_ISL_1583188, EPI_ISL_1583252, EPI_ISL_1583253, EPI_ISL_1585269, EPI_ISL_1585273, EPI_ISL_1585940, EPI_ISL_1585942 | Armed Forces Institute of Pathology (AFIP), Dhaka Cantonment | Genomic Research Lab, BCSIR | Md. Murshed Hasan Sarkar, Abu Sayeed Mohammad Mahmud, Mohammad Samir Uzzaman, Eshrar Osman, Md. Ahasan Habib, Shahina Akter, Tanjina Akhter Banu, Barna Goswami, Iffat Jahan, Md. Saddam Hossain, Mohammad Mohi Uddin, Md. Kamrul Islam, Mohammad Mizanur Rahman, Susane Giti, Md. Salim Khan |
| EPI_ISL_1587429, EPI_ISL_1587430, EPI_ISL_1587431, EPI_ISL_1587432, EPI_ISL_1587433, EPI_ISL_1587434, EPI_ISL_1587435, EPI_ISL_1587436, EPI_ISL_1587437, EPI_ISL_1587438, EPI_ISL_1587439, EPI_ISL_1587440, EPI_ISL_1587441, EPI_ISL_1587442, EPI_ISL_1587443, EPI_ISL_1587444, EPI_ISL_1587445, EPI_ISL_1587446, EPI_ISL_1587447 | | | |
| see above | Institute for Developing Science and Health Initiatives (ideSHi) | Institute for Developing Science and Health Initiatives (ideSHi) | Hassan Afrad, Sadia Rahman, Fidausi Qadri, Tahmina Shirin |
| EPI_ISL_1588437, EPI_ISL_1588593 | Armed Forces Institute of Pathology (AFIP), Dhaka Cantonment | Genomic Research Lab, BCSIR | Abu Sayeed Mohammad Mahmud, Md. Murshed Hasan Sarkar, Mohammad Samir Uzzaman, Eshrar Osman, Md. Ahasan Habib, Shahina Akter, Tanjina Akhter Banu, Barna Goswami, Iffat Jahan, Md. Saddam Hossain, Mohammad Mohi Uddin, Md. Kamrul Islam, Mohammad Mizanur Rahman, Susane Giti, Md. Salim Khan |
| EPI_ISL_1593804 | Armed Forces Institute of Pathology (AFIP), Dhaka Cantonment | Genomic Research Lab, BCSIR | Abu Sayeed Mohammad Mahmud, Md. Murshed Hasan Sarkar, Mohammad Samir Uzzaman, Eshrar Osman, Md. Ahasan Habib, Shahina Akter, Tanjina Akhter Banu, Barna Goswami, Iffat Jahan, Md. Saddam Hossain, Mohammad Mohi Uddin, Md. Kamrul Islam, Mohammad Mizanur Rahman, Susane Giti, Md. Salim Khan |
| EPI_ISL_1593853, EPI_ISL_1593854, EPI_ISL_1593855 | Institute for Developing Science and Health Initiatives (ideSHi) | Institute for Developing Science and Health Initiatives (ideSHi) | Hassan Afrad, Sadia Rahman, Fidausi Qadri, Tahmina Shirin |
| EPI_ISL_1593856, EPI_ISL_1593857, EPI_ISL_1593858 | Armed Forces Institute of Pathology (AFIP), Dhaka Cantonment | Genomic Research Lab, BCSIR | Mohammad Mizanur Rahman, Md. Murshed Hasan Sarkar, Mohammad Samir Uzzaman, Eshrar Osman, Md. Ahasan Habib, Shahina Akter, Tanjina Akhter Banu, Abu Sayeed Mohammad Mahmud, Barna Goswami, Iffat Jahan, Md. Saddam Hossain, Mohammad Mohi Uddin, Md. Kamrul Islam, Susane Giti, Md. Salim Khan |
| EPI_ISL_1593859, EPI_ISL_1593860, EPI_ISL_1593861, EPI_ISL_1593862 | Armed Forces Institute of Pathology (AFIP), Dhaka Cantonment | Genomic Research Lab, BCSIR | Susane Giti, Md. Murshed Hasan Sarkar, Mohammad Samir Uzzaman, Eshrar Osman, Md. Ahasan Habib, Shahina Akter, Tanjina Akhter Banu, Abu Sayeed Mohammad Mahmud, Barna Goswami, Iffat Jahan, Md. Saddam Hossain, Mohammad Mohi Uddin, Md. Kamrul Islam, Mohammad Mizanur Rahman, Md. Salim Khan |
| EPI_ISL_1593863, EPI_ISL_1593879 | Armed Forces Institute of Pathology (AFIP), Dhaka Cantonment | Genomic Research Lab, BCSIR | Shahina Akter, Md. Murshed Hasan Sarkar, Mohammad Samir Uzzaman, Eshrar Osman, Md. Ahasan Habib, Tanjina Akhter Banu, Abu Sayeed Mohammad Mahmud, Barna Goswami, Iffat Jahan, Md. Saddam Hossain, Mohammad Mohi Uddin, Md. Kamrul Islam, Mohammad Mizanur Rahman, Susane Giti, Md. Salim Khan |
| EPI_ISL_1593880, EPI_ISL_1593888 | Armed Forces Institute of Pathology (AFIP), Dhaka Cantonment | Genomic Research Lab, BCSIR | Tanjina Akhter Banu, Md. Murshed Hasan Sarkar, Mohammad Samir Uzzaman, Eshrar Osman, Md. Ahasan Habib, Shahina Akter, Tanjina Akhter Banu, Abu Sayeed Mohammad Mahmud, Barna Goswami, Iffat Jahan, Md. Saddam Hossain, Mohammad Mohi Uddin, Md. Kamrul Islam, Mohammad Mizanur Rahman, Susane Giti, Md. Salim Khan |
| EPI_ISL_1593894, EPI_ISL_1595718, EPI_ISL_1595721, EPI_ISL_1595831 | Armed Forces Institute of Pathology (AFIP), Dhaka Cantonment | Genomic Research Lab, BCSIR | Md. Murshed Hasan Sarkar, Mohammad Samir Uzzaman, Eshrar Osman, Md. Ahasan Habib, Shahina Akter, Tanjina Akhter Banu, Abu Sayeed Mohammad Mahmud, Barna Goswami, Iffat Jahan, Md. Saddam Hossain, Mohammad Mohi Uddin, Md. Kamrul Islam, Mohammad Mizanur Rahman, Susane Giti, Md. Salim Khan |
| EPI_ISL_1595832, EPI_ISL_1595833, EPI_ISL_1595834 | Armed Forces Institute of Pathology (AFIP), Dhaka Cantonment | Genomic Research Lab, BCSIR | Barna Goswami, Md. Murshed Hasan Sarkar, Mohammad Samir Uzzaman, Eshrar Osman, Md. Ahasan Habib, Shahina Akter, Tanjina Akhter Banu, Abu Sayeed Mohammad Mahmud, Iffat Jahan, Md. Saddam Hossain, Mohammad Mohi Uddin, Md. Kamrul Islam, Mohammad Mizanur Rahman, Susane Giti, Md. Salim Khan |
| EPI_ISL_1595835, EPI_ISL_1595837 | Armed Forces Institute of Pathology (AFIP), Dhaka Cantonment | Genomic Research Lab, BCSIR | Iffat Jahan, Md. Murshed Hasan Sarkar, Mohammad Samir Uzzaman, Eshrar Osman, Md. Ahasan Habib, Shahina Akter, Tanjina Akhter Banu, Abu Sayeed Mohammad Mahmud, Barna Goswami, Md. Saddam Hossain, Mohammad Mohi Uddin, Md. Kamrul Islam, Mohammad Mizanur Rahman, Susane Giti, Md. Salim Khan |

| | | | |
|---|---|---|---|
| EPI_ISL_1595839, EPI_ISL_1595840 | Armed Forces Institute of Pathology (AFIP), Dhaka Cantonment | Genomic Research Lab, BCSIR | Md. Saddam Hossain, Md. Murshed Hasan Sarkar, Mohammad Samir Uzzaman, Eshrar Osman, Md. Ahasan Habib, Shahina Akter, Tanjina Akhter Banu, Abu Sayeed Mohammad Mahmud, Barna Goswami, Iffat Jahan, Mohammad Mohi Uddin, Md. Kamrul Islam, Mohammad Mizanur Rahman, Susane Giti, Md. Salim Khan |
| EPI_ISL_1595841, EPI_ISL_1595842 | Armed Forces Institute of Pathology (AFIP), Dhaka Cantonment | Genomic Research Lab, BCSIR | Mohammad Mohi Uddin, Md. Murshed Hasan Sarkar, Mohammad Samir Uzzaman, Eshrar Osman, Md. Ahasan Habib, Shahina Akter, Tanjina Akhter Banu, Abu Sayeed Mohammad Mahmud, Barna Goswami, Iffat Jahan, Md. Saddam Hossain, Md. Kamrul Islam, Mohammad Mizanur Rahman, Susane Giti, Md. Salim Khan |
| EPI_ISL_1595843 | Armed Forces Institute of Pathology (AFIP), Dhaka Cantonment | Genomic Research Lab, BCSIR | Md. Murshed Hasan Sarkar, Mohammad Samir Uzzaman, Eshrar Osman, Md. Ahasan Habib, Shahina Akter, Tanjina Akhter Banu, Abu Sayeed Mohammad Mahmud, Barna Goswami, Iffat Jahan, Md. Saddam Hossain, Mohammad Mohi Uddin, Md. Kamrul Islam, Mohammad Mizanur Rahman, Susane Giti, Md. Salim Khan |
| EPI_ISL_1615663, EPI_ISL_1615664 | Armed Forces Institute of Pathology (AFIP), Dhaka Cantonment | Genomic Research Lab, BCSIR | Mohammad Mizanur Rahman, Md. Murshed Hasan Sarkar, Mohammad Samir Uzzaman, Eshrar Osman, Md. Ahasan Habib, Shahina Akter, Tanjina Akhter Banu, Abu Sayeed Mohammad Mahmud, Barna Goswami, Iffat Jahan, Md. Saddam Hossain, Mohammad Mohi Uddin, Md. Kamrul Islam, Susane Giti, Md. Salim Khan |
| EPI_ISL_1616000, EPI_ISL_1616606 | Armed Forces Institute of Pathology (AFIP), Dhaka Cantonment | Genomic Research Lab, BCSIR | Susane Giti, Md. Murshed Hasan Sarkar, Mohammad Samir Uzzaman, Eshrar Osman, Md. Ahasan Habib, Shahina Akter, Tanjina Akhter Banu, Abu Sayeed Mohammad Mahmud, Barna Goswami, Iffat Jahan, Md. Saddam Hossain, Mohammad Mohi Uddin, Md. Kamrul Islam, Mohammad Mizanur Rahman, Md. Salim Khan |
| EPI_ISL_1616608, EPI_ISL_1616615, EPI_ISL_1616616, EPI_ISL_1616835, EPI_ISL_1616836, EPI_ISL_1616838 | Armed Forces Institute of Pathology (AFIP), Dhaka Cantonment | Genomic Research Lab, BCSIR | Md. Murshed Hasan Sarkar, Mohammad Samir Uzzaman, Eshrar Osman, Md. Ahasan Habib, Shahina Akter, Tanjina Akhter Banu, Abu Sayeed Mohammad Mahmud, Barna Goswami, Iffat Jahan, Md. Saddam Hossain, Mohammad Mohi Uddin, Md. Kamrul Islam, Mohammad Mizanur Rahman, Susane Giti, Md. Salim Khan |
| EPI_ISL_1629810, EPI_ISL_1629811 | Armed Forces Institute of Pathology (AFIP), Dhaka Cantonment | Genomic Research Lab, BCSIR | Md. Murshed Hasan Sarkar, Mohammad Samir Uzzaman, Eshrar Osman, Md. Ahasan Habib, Shahina Akter, Tanjina Akhter Banu, Abu Sayeed Mohammad Mahmud, Barna Goswami, Iffat Jahan, Md. Saddam Hossain, Mohammad Mohi Uddin, Md. Kamrul Islam, Mohammad Mizanur Rahman, Susane Giti, Md. Salim Khan |
| EPI_ISL_1634445, EPI_ISL_1634446, EPI_ISL_1634447, EPI_ISL_1634448, EPI_ISL_1634449, EPI_ISL_1634450, EPI_ISL_1634451, EPI_ISL_1634452, EPI_ISL_1634453, EPI_ISL_1634455, EPI_ISL_1634456, EPI_ISL_1634457, EPI_ISL_1634458, EPI_ISL_1634459, EPI_ISL_1634460, EPI_ISL_1634461, EPI_ISL_1634462, EPI_ISL_1634463 | | | |
| see above | Child Health Research Foundation | Child Health Research Foundation | CHRF Bangladesh Genomics Team |
| EPI_ISL_1636523, EPI_ISL_1653815 | DNA Solution Ltd. | Genomic Research Lab, BCSIR | Md. Murshed Hasan Sarkar, Mohammad Samir Uzzaman, Eshrar Osman, Md. Ahasan Habib, Shahina Akter, Tanjina Akhter Banu, Abu Sayeed Mohammad Mahmud, Barna Goswami, Iffat Jahan, Md. Saddam Hossain, Mohammad Mohi Uddin, Mohammad Fazle Alam Rabbi, Md Firoz Kabir, Kazi Nadim Hasan, Md. Mizanur Rahman, Md. Abdul Khaleque, Sharif Akhteruzzamani, Md. Salim Khan |
| EPI_ISL_1653893, EPI_ISL_1653894, EPI_ISL_1653920, EPI_ISL_1653921, EPI_ISL_1653922 | DNA Solution Ltd. | Genomic Research Lab, BCSIR | Mohammad Fazle Alam Rabbi, Md. Murshed Hasan Sarkar, Mohammad Samir Uzzaman, Eshrar Osman, Md. Ahasan Habib, Shahina Akter, Tanjina Akhter Banu, Abu Sayeed Mohammad Mahmud, Barna Goswami, Iffat Jahan, Md. Saddam Hossain, Mohammad Mohi Uddin, Md Firoz Kabir, Kazi Nadim Hasan, Md. Mizanur Rahman, Md. Abdul Khaleque, Sharif Akhteruzzamani, Md. Salim Khan |
| EPI_ISL_1653926 | DNA Solution Ltd. | Genomic Research Lab, BCSIR | Abu Sayeed Mohammad Mahmud, Md. Murshed Hasan Sarkar, Mohammad Samir Uzzaman, Eshrar Osman, Md. Ahasan Habib, Shahina Akter, Tanjina Akhter Banu, Barna Goswami, Iffat Jahan, Md. Saddam Hossain, Mohammad Mohi Uddin, Mohammad Fazle Alam Rabbi, Md Firoz Kabir, Kazi Nadim Hasan, Md. Mizanur Rahman, Md. Abdul Khaleque, Sharif Akhteruzzamani, Md. Salim Khan |
| EPI_ISL_1657084, EPI_ISL_1657085, EPI_ISL_1657086, EPI_ISL_1657087, EPI_ISL_1657088, EPI_ISL_1669903, EPI_ISL_1669904, EPI_ISL_1669905, EPI_ISL_1669906, EPI_ISL_1669907 | Institute for Developing Science and Health Initiatives (ideSHi) | Institute for Developing Science and Health Initiatives (ideSHi) | Hassan Afrad, Sadia Rahman, Fidausi Qadri, Tahmina Shirin |
| EPI_ISL_1673289 | DNA Solution Ltd. | Genomic Research Lab, BCSIR | Barna Goswami, Md. Murshed Hasan Sarkar, Mohammad Samir Uzzaman, Eshrar Osman, Md. Ahasan Habib, Shahina Akter, Tanjina Akhter Banu, Abu Sayeed Mohammad MahmudIffat Jahan, Md. Saddam Hossain, Mohammad Mohi Uddin, Mohammad Fazle Alam Rabbi, Md Firoz Kabir, Kazi Nadim Hasan, Md. Mizanur Rahman, Md. Abdul Khaleque, Sharif Akhteruzzamani, Md. Salim Khan |
| EPI_ISL_1673292 | DNA Solution Ltd. | Genomic Research Lab, BCSIR | Iffat Jahan, Md. Murshed Hasan Sarkar, Mohammad Samir Uzzaman, Eshrar Osman, Md. Ahasan Habib, Shahina Akter, Tanjina Akhter Banu, Abu Sayeed Mohammad Mahmud, Barna Goswami, Md. Saddam Hossain, Mohammad Mohi Uddin, Mohammad Fazle Alam Rabbi, Md Firoz Kabir, Kazi Nadim Hasan, Md. Mizanur Rahman, Md. Abdul Khaleque, Sharif Akhteruzzamani, Md. Salim Khan |
| EPI_ISL_1673302 | DNA Solution Ltd. | Genomic Research Lab, BCSIR | Md. Saddam Hossain, Md. Murshed Hasan Sarkar, Mohammad Samir Uzzaman, Eshrar Osman, Md. Ahasan Habib, Shahina Akter, Tanjina Akhter Banu, Abu Sayeed Mohammad Mahmud, Barna Goswami, Iffat Jahan, Mohammad Mohi Uddin, Mohammad Fazle Alam Rabbi, Md Firoz Kabir, Kazi Nadim Hasan, Md. Mizanur Rahman, Md. Abdul Khaleque, Sharif Akhteruzzamani, Md. Salim Khan |
| EPI_ISL_1673334, EPI_ISL_1673339, EPI_ISL_1673343, EPI_ISL_1673415 | DNA Solution Ltd. | Genomic Research Lab, BCSIR | Md. Murshed Hasan Sarkar, Mohammad Samir Uzzaman, Eshrar Osman, Md. Ahasan Habib, Shahina Akter, Tanjina Akhter Banu, Abu Sayeed Mohammad Mahmud, Barna Goswami, Iffat Jahan, Md. Saddam Hossain, Mohammad Mohi Uddin, Mohammad Fazle Alam Rabbi, Md Firoz Kabir, Kazi Nadim Hasan, Md. Mizanur Rahman, Md. Abdul Khaleque, Sharif Akhteruzzamani, Md. Salim Khan |
| EPI_ISL_1673673 | Genomic Research Lab, BCSIR | Genomic Research Lab, BCSIR | Tanjina Akhter Banu, Md. Murshed Hasan Sarkar, Mohammad Samir Uzzaman, Eshrar Osman, Md. Ahasan Habib, Shahina Akter, Abu Sayeed Mohammad Mahmud, Barna Goswami, Iffat Jahan, Md. Saddam Hossain, Mohammad Mohi Uddin, Mohammad Fazle Alam Rabbi, Md Firoz Kabir, Kazi Nadim Hasan, Md. Mizanur Rahman, Md. Abdul Khaleque, Sharif Akhteruzzamani, Md. Salim Khan |
| EPI_ISL_1673680 | Genomic Research Lab, BCSIR | Genomic Research Lab, BCSIR | Tanjina Akhter Banu, Md. Murshed Hasan Sarkar, Mohammad Samir Uzzaman, Eshrar Osman, Md. Ahasan Habib, Shahina Akter, Abu Sayeed Mohammad Mahmud, Barna Goswami, Iffat Jahan, Md. Saddam Hossain, Mohammad Mohi Uddin, Md. Salim Khan |
| EPI_ISL_1714800, EPI_ISL_1714801, EPI_ISL_1714805, EPI_ISL_1714806 | GRMDCR & JU | Genomic Research Lab, BCSIR | Md. Murshed Hasan Sarkar, Mohammad Samir Uzzaman, Eshrar Osman, Md. Ahasan Habib, Shahina Akter, Tanjina Akhter Banu, Abu Sayeed Mohammad Mahmud, Barna Goswami, Iffat Jahan, Md. Saddam Hossain, Mohammad Mohi Uddin, Nihad Adnan, Mohd Raeed Jamiruddin, M Ahsanul Haq, Mohib Ullah Khondoker, Maha Jamiruddin, Md. Rubel Hossain, Nowshin Jahan, Tamanna Ali, Shahad Saif Khandker, M Firoz Ahmed, Md. Salim Khan |
| EPI_ISL_1715160, EPI_ISL_1715161, EPI_ISL_1715162, EPI_ISL_1715163, EPI_ISL_1715164, EPI_ISL_1715165, EPI_ISL_1715166, EPI_ISL_1715167, EPI_ISL_1715168, EPI_ISL_1715169, EPI_ISL_1715170, EPI_ISL_1717025 | | | |
| see above | Virology Laboratory, International Centre for Diarrhoeal Disease Research, Bangladesh (ICDDR,B) | Virology Laboratory, International Centre for Diarrhoeal Disease Research, Bangladesh (ICDDR,B) | Mohammad Enayet Hossain, Mojnu Miah, Rashedul Hasan, Md. Mahfuzur Rahman, Mohammed Ziaur Rahman, Mustafizur Rahman |
| EPI_ISL_1719905, EPI_ISL_1719907 | GRMDCR & JU | Genomic Research Lab, BCSIR | Shahina Akter, Md. Murshed Hasan Sarkar, Mohammad Samir Uzzaman, Eshrar Osman, Md. Ahasan Habib, Tanjina Akhter Banu, Abu Sayeed Mohammad Mahmud, Barna Goswami, Iffat Jahan, Md. Saddam Hossain, Mohammad Mohi Uddin, Nihad Adnan, Mohd Raeed Jamiruddin, M Ahsanul Haq, Mohib Ullah Khondoker, Maha Jamiruddin, Md. Rubel Hossain, Nowshin Jahan, Tamanna Ali, Shahad Saif Khandker, M Firoz Ahmed, Md. Salim Khan |
| EPI_ISL_1719908 | GRMDCR & JU | Genomic Research Lab, BCSIR | Tanjina Akhter Banu, Md. Murshed Hasan Sarkar, Mohammad Samir Uzzaman, Eshrar Osman, Md. Ahasan Habib, Shahina Akter, Abu Sayeed Mohammad Mahmud, Barna Goswami, Iffat Jahan, Md. Saddam Hossain, Mohammad Mohi Uddin, Nihad Adnan, Mohd Raeed Jamiruddin, M Ahsanul Haq, Mohib Ullah Khondoker, Maha Jamiruddin, Md. Rubel Hossain, Nowshin Jahan, Tamanna Ali, Shahad Saif Khandker, M Firoz Ahmed, Md. Salim Khan |
| EPI_ISL_1719916 | GRMDCR & JU | Genomic Research Lab, BCSIR | Tanjina Akhter Banu, Md. Murshed Hasan Sarkar, Mohammad Samir Uzzaman, Eshrar Osman, Md. Ahasan Habib, Shahina Akter, Abu Sayeed Mohammad Mahmud, Barna Goswami, Iffat Jahan, Md. Saddam Hossain, Mohammad Mohi Uddin, Md. Kamrul Islam, Nihad Adnan, Mohd Raeed Jamiruddin, M Ahsanul Haq, Mohib Ullah Khondoker, Maha Jamiruddin, Md. Rubel Hossain, Nowshin Jahan, Tamanna Ali, Shahad Saif Khandker, M Firoz Ahmed, Md. Salim Khan |
| EPI_ISL_1720931, EPI_ISL_1721346 | GRMDCR & JU | Genomic Research Lab, BCSIR | Abu Sayeed Mohammad Mahmud, Md. Murshed Hasan Sarkar, Mohammad Samir Uzzaman, Eshrar Osman, Md. Ahasan Habib, Shahina Akter, Tanjina |

| | | | |
|---|---|---|---|
| | | | Akhter Banu, Barna Goswami, Iffat Jahan, Md. Saddam Hossain, Mohammad Mohi Uddin, Md. Kamrul Islam, Nihad Adnan, Mohd Raeed Jamiruddin, M Ahsanul Haq, Mohib Ullah Khondoker, Maha Jamiruddin, Md. Rubel Hossain, Nowshin Jahan, Tamanna Ali, Shahad Saif Khandker, M Firoz Ahmed, Md. Salim Khan |
| EPI_ISL_1721825, EPI_ISL_1722306 | GRMDCR & JU | Genomic Research Lab, BCSIR | Barna Goswami, Md. Murshed Hasan Sarkar, Mohammad Samir Uzzaman, Eshrar Osman, Md. Ahasan Habib, Shahina Akter, Tanjina Akhter Banu, Abu Sayeed Mohammad Mahmud, Iffat Jahan, Md. Saddam Hossain, Mohammad Mohi Uddin, Md. Kamrul Islam, Nihad Adnan, Mohd Raeed Jamiruddin, M Ahsanul Haq, Mohib Ullah Khondoker, Maha Jamiruddin, Md. Rubel Hossain, Nowshin Jahan, Tamanna Ali, Shahad Saif Khandker, M Firoz Ahmed, Md. Salim Khan |
| EPI_ISL_1722681 | GRMDCR & JU | Genomic Research Lab, BCSIR | Iffat Jahan, Md. Murshed Hasan Sarkar, Mohammad Samir Uzzaman, Eshrar Osman, Md. Ahasan Habib, Shahina Akter, Tanjina Akhter Banu, Abu Sayeed Mohammad Mahmud, Barna Goswami, Md. Saddam Hossain, Mohammad Mohi Uddin, Md. Kamrul Islam, Nihad Adnan, Mohd Raeed Jamiruddin, M Ahsanul Haq, Mohib Ullah Khondoker, Maha Jamiruddin, Md. Rubel Hossain, Nowshin Jahan, Tamanna Ali, Shahad Saif Khandker, M Firoz Ahmed, Md. Salim Khan |
| EPI_ISL_1742836 | GRMDCR & JU | Genomic Research Lab, BCSIR | Md. Saddam Hossain, Md. Murshed Hasan Sarkar, Mohammad Samir Uzzaman, Eshrar Osman, Md. Ahasan Habib, Shahina Akter, Tanjina Akhter Banu, Abu Sayeed Mohammad Mahmud, Barna Goswami, Iffat Jahan, Mohammad Mohi Uddin, Md. Kamrul Islam, Nihad Adnan, Mohd Raeed Jamiruddin, M Ahsanul Haq, Mohib Ullah Khondoker, Maha Jamiruddin, Md. Rubel Hossain, Nowshin Jahan, Tamanna Ali, Shahad Saif Khandker, M Firoz Ahmed, Md. Salim Khan |
| EPI_ISL_1742837, EPI_ISL_1742838, EPI_ISL_1743263, EPI_ISL_1743591 | GRMDCR & JU | Genomic Research Lab, BCSIR | Md. Murshed Hasan Sarkar, Mohammad Samir Uzzaman, Eshrar Osman, Md. Ahasan Habib, Shahina Akter, Tanjina Akhter Banu, Abu Sayeed Mohammad Mahmud, Barna Goswami, Iffat Jahan, Md. Saddam Hossain, Mohammad Mohi Uddin, Md. Kamrul Islam, Nihad Adnan, Mohd Raeed Jamiruddin, M Ahsanul Haq, Mohib Ullah Khondoker, Maha Jamiruddin, Md. Rubel Hossain, Nowshin Jahan, Tamanna Ali, Shahad Saif Khandker, M Firoz Ahmed, Md. Salim Khan |
| EPI_ISL_1750956, EPI_ISL_1750957, EPI_ISL_1750958, EPI_ISL_1750959, EPI_ISL_1750960, EPI_ISL_1750961, EPI_ISL_1750962 | Institute of Epidemiology, Disease Control and Research (IEDCR) | Institute for Developing Science and Health Initiatives (ideSHi) | Hassan Afrad, Sadia Rahman, Fidausi Qadri, Tahmina Shirin |
| EPI_ISL_1752689, EPI_ISL_1752690, EPI_ISL_1752691, EPI_ISL_1752692, EPI_ISL_1752693, EPI_ISL_1752694, EPI_ISL_1752702, EPI_ISL_1752703, EPI_ISL_1752704, EPI_ISL_1752705, EPI_ISL_1752706, EPI_ISL_1752707 | | | |
| see above | Child Health Research Foundation | Child Health Research Foundation | CHRF Bangladesh Genomics Team |
| EPI_ISL_1760648, EPI_ISL_1760649, EPI_ISL_1760650 | Institute of Epidemiology, Disease Control and Research (IEDCR) | Institute for Developing Science and Health Initiatives (ideSHi) | Hassan Afrad, Sadia Rahman, Fidausi Qadri, Tahmina Shirin |
| EPI_ISL_1790209 | Department of Genetic Engineering and Biotechnology, Shahjalal University of Science and Technology | Genomic Research Lab, BCSIR | Md. Murshed Hasan Sarkar, Abu Sayeed Mohammad Mahmud, Mohammad Samir Uzzaman, Eshrar Osman, Md. Ahasan Habib, Shahina Akter, Tanjina Akhter Banu, Barna Goswami, Iffat Jahan, Md. Saddam Hossain, Mohammad Mohi Uddin, Md. Kamrul Islam, Md. Shamsul Haque Prodhan, Md. Hammadul Hoque, G. M. Nurnabi Azad Jewel, Md. Nazmul Hasan, Md. Fahmid Hossain Bhuiyan, Md. Asraful Jahan, Ajit Ghosh, Md. Akkas Ali, Md. Salim Khan |
| EPI_ISL_1790213 | Department of Genetic Engineering and Biotechnology, Shahjalal University of Science and Technology | Genomic Research Lab, BCSIR | Tanjina Akhter Banu, Md. Murshed Hasan Sarkar, Abu Sayeed Mohammad Mahmud, Mohammad Samir Uzzaman, Eshrar Osman, Md. Ahasan Habib, Shahina Akter, Barna Goswami, Iffat Jahan, Md. Saddam Hossain, Mohammad Mohi Uddin, Md. Kamrul Islam, Md. Shamsul Haque Prodhan, Md. Hammadul Hoque, G. M. Nurnabi Azad Jewel, Md. Nazmul Hasan, Md. Fahmid Hossain Bhuiyan, Md. Asraful Jahan, Ajit Ghosh, Md. Akkas Ali, Md. Salim Khan |
| EPI_ISL_1793784, EPI_ISL_1793785, EPI_ISL_1793786, EPI_ISL_1793787, EPI_ISL_1793788, EPI_ISL_1793789, EPI_ISL_1793790, EPI_ISL_1793791 | Virology Laboratory, International Centre for Diarrhoeal Disease Research, Bangladesh (ICDDR,B) | Virology Laboratory, International Centre for Diarrhoeal Disease Research, Bangladesh (ICDDR,B) | Mohammad Enayet Hossain, Mojnu Miah, Rashedul Hasan, Md. Mahfuzur Rahman, Mohammed Ziaur Rahman, Mustafizur Rahman |
| EPI_ISL_1805696 | Department of Genetic Engineering and Biotechnology, Shahjalal University of Science and Technology | Genomic Research Lab, BCSIR | Md. Murshed Hasan Sarkar, Abu Sayeed Mohammad Mahmud, Mohammad Samir Uzzaman, Eshrar Osman, Md. Ahasan Habib, Shahina Akter, Tanjina Akhter Banu, Barna Goswami, Iffat Jahan, Md. Saddam Hossain, Mohammad Mohi Uddin, Md. Kamrul Islam, Md. Shamsul Haque Prodhan, Md. Hammadul Hoque, G. M. Nurnabi Azad Jewel, Md. Nazmul Hasan, Md. Fahmid Hossain Bhuiyan, Md. Asraful Jahan, Ajit Ghosh, Md. Akkas Ali, Md. Salim Khan |
| EPI_ISL_1805701 | Genomic Research Lab, BCSIR | Genomic Research Lab, BCSIR | Md. Salim Khan, Md. Murshed Hasan Sarkar, Abu Sayeed Mohammad Mahmud, Mohammad Samir Uzzaman, Eshrar Osman, Md. Ahasan Habib, Shahina Akter, Tanjina Akhter Banu, Barna Goswami, Iffat Jahan, Md. Saddam Hossain, Mohammad Mohi Uddin, Md. Kamrul Islam, Md. Shamsul Haque Prodhan, Md. Hammadul Hoque, G. M. Nurnabi Azad Jewel, Md. Nazmul Hasan, Md. Fahmid Hossain Bhuiyan, Md. Asraful Jahan, Ajit Ghosh, Md. Akkas Ali, Md. Salim Khan |
| EPI_ISL_1825676, EPI_ISL_1825678, EPI_ISL_1825679, EPI_ISL_1825680, EPI_ISL_1825681, EPI_ISL_1825682, EPI_ISL_1825683, EPI_ISL_1825684, EPI_ISL_1825685, EPI_ISL_1825686 | COVID-19 Detection Lab, Chattogram Veterinary and Animal Sciences University | Genomic Research Lab, Bangladesh Council of Scientific and Industrial Research | Goutam Buddha Das, Tridip Das, Tanvir Ahmad Nizami, Eaftekhar Ahmed Rana, Md. Sirazul Islam, Pronesh Dutta, Sharmin Chowdhury, Md. Morshed Hasan Sarkar, Md. Salim Khan, Paritosh Kumar Biswas |
| EPI_ISL_1828779, EPI_ISL_1828780, EPI_ISL_1828781, EPI_ISL_1828782 | Virology Laboratory, International Centre for Diarrhoeal Disease Research, Bangladesh (ICDDR,B) | Virology Laboratory, International Centre for Diarrhoeal Disease Research, Bangladesh (ICDDR,B) | Mohammad Enayet Hossain, Mojnu Miah, Rashedul Hasan, Md. Mahfuzur Rahman, Mohammed Ziaur Rahman, Mustafizur Rahman |
| EPI_ISL_1915108, EPI_ISL_1915109, EPI_ISL_1915110, EPI_ISL_1915111, EPI_ISL_1915112, EPI_ISL_1915113, EPI_ISL_1915114, EPI_ISL_1915115, EPI_ISL_1915116, EPI_ISL_1915117, EPI_ISL_1915118, EPI_ISL_1915119, EPI_ISL_1915120 | | | |
| see above | Institute for Developing Science and Health Initiatives (ideSHi) | Institute for Developing Science and Health Initiatives (ideSHi) | Hassan Afrad, Sadia Rahman, Fidausi Qadri, Tahmina Shirin |
| EPI_ISL_1915436, EPI_ISL_1915437, EPI_ISL_1915438, EPI_ISL_1915439 | Institute of Epidemiology, Disease Control and Research (IEDCR) | Institute for Developing Science and Health Initiatives (ideSHi) | Hassan Afrad, Sadia Rahman, Fidausi Qadri, Tahmina Shirin |
| EPI_ISL_1920962 | Institute for Developing Science and Health Initiatives (ideSHi) | Institute for Developing Science and Health Initiatives (ideSHi) | Hassan Afrad, Sadia Rahman, Fidausi Qadri, Tahmina Shirin |
| EPI_ISL_1938475, EPI_ISL_1938476, EPI_ISL_1938477 | Institute of Epidemiology, Disease Control and Research (IEDCR) | Institute for Developing Science and Health Initiatives (ideSHi) | Hassan Afrad, Sadia Rahman, Fidausi Qadri, Tahmina Shirin |
| EPI_ISL_1942249 | Genome Centre | Genome Centre | Md. Shazid Hasan, Shovon Lal Sarkar, Ali Ahsan Setu, Tanay Chakrabarty, A. S. M. Rubayet Ul Alam, M. Shaminur Rahman, Ovinu Kibria Islam, M. Tanvir Islam, Hassan M. Al-Emran, Iqbal Kabir Jahid, M. Anwar Hossain |
| EPI_ISL_2002482 | Genomic Research Lab, BCSIR | Genomic Research Lab, BCSIR | Md. Murshed Hasan Sarkar, Abu Sayeed Mohammad Mahmud, Mohammad Samir Uzzaman, Eshrar Osman, Md. Ahasan Habib, Shahina Akter, Tanjina Akhter Banu, Barna Goswami, Iffat Jahan, Md. Saddam Hossain, Mohammad Mohi Uddin, Md. Kamrul Islam, Md. Salim Khan |
| EPI_ISL_2003459 | Virology Laboratory, IDD, icddr,b | Virology Laboratory, IDD, icddr,b | Muntasir Alam, M Ishrat Jahan, Shafina Jahan, Rubaiyat Yameen, Afruna Rahman, Emily S. Gurley, Shams El Arifeen, Mustafizur Rahman |
| EPI_ISL_2036272 | Genome Centre | Genome Centre | Shovon Lal Sarkar, Md. Shazid Hasan, Ali Ahsan Setu, Prosanto Kumar Das, A. S. M. Rubayet Ul Alam, Ovinu Kibria Islam, Md. Tanvir Islam, Hassan Md. Al-Emran, Iqbal Kabir Jahid, M. Anwar Hossain |
| EPI_ISL_2037865, EPI_ISL_2038623, EPI_ISL_2038944, EPI_ISL_2039394, EPI_ISL_2039757, EPI_ISL_2040202, EPI_ISL_2043212, EPI_ISL_2043671, EPI_ISL_2044075, EPI_ISL_2044525, EPI_ISL_2044950, EPI_ISL_2045661, EPI_ISL_2046273, EPI_ISL_2046750, EPI_ISL_2047053 | | | |
| see above | Biological Hazard and Health Research Laboratory, Centre for Advanced Research in Science (CARS), University of Dhaka | Genomic Research Lab, BCSIR | Md Mizanur Rahaman, Md Abdul Malek, Md. Shaminur Rahman, Latiful Bari, Md. Murshed Hasan Sarkar, Abu Sayeed Mohammad Mahmud, Mohammad Samir Uzzaman, Eshrar Osman, Md. Ahasan Habib, Shahina Akter, Tanjina Akhter Banu, Barna Goswami, Iffat Jahan, Md. Saddam Hossain, Mohammad Mohi Uddin, Md. Kamrul Islam, Md. Salim Khan |
| EPI_ISL_2105565 | Institute of Epidemiology, Disease Control and Research (IEDCR) | ideSHi-IEDCR-icddr,b | Hassan Afrad, Sadia Rahman, Manjur Hossain Khan, Firdausi Qadri, Tahmina Shirin |

| | | | |
|---|---|---|---|
| EPI_ISL_2178947, EPI_ISL_2178948, EPI_ISL_2178949, EPI_ISL_2178950 | Praava Health | Child Health Research Foundation | CHRF Bangladesh Genomics Team, Zaheed Husain, Shafiul Azam |
| EPI_ISL_2178951 | Child Health Research Foundation | Child Health Research Foundation | CHRF Bangladesh Genomics Team |
| EPI_ISL_2178952, EPI_ISL_2178953, EPI_ISL_2178954 | Praava Health | Child Health Research Foundation | CHRF Bangladesh Genomics Team, Zaheed Husain, Shafiul Azam |
| EPI_ISL_2178955, EPI_ISL_2178956, EPI_ISL_2178957 | Child Health Research Foundation | Child Health Research Foundation | CHRF Bangladesh Genomics Team |
| EPI_ISL_2178958 | Praava Health | Child Health Research Foundation | CHRF Bangladesh Genomics Team, Zaheed Husain, Shafiul Azam |
| EPI_ISL_2178959 | Child Health Research Foundation | Child Health Research Foundation | CHRF Bangladesh Genomics Team |
| EPI_ISL_2178960, EPI_ISL_2178961, EPI_ISL_2178962, EPI_ISL_2178963, EPI_ISL_2178964, EPI_ISL_2178965, EPI_ISL_2178966, EPI_ISL_2178967, EPI_ISL_2178968, EPI_ISL_2178969, EPI_ISL_2178970, EPI_ISL_2178971, EPI_ISL_2178972, EPI_ISL_2178973, EPI_ISL_2178974, EPI_ISL_2178975 | | | |
| see above | Praava Health | Child Health Research Foundation | CHRF Bangladesh Genomics Team, Zaheed Husain, Shafiul Azam |
| EPI_ISL_2179666, EPI_ISL_2179667 | Institute for Developing Science and Health Initiatives (ideSHi) | Institute for Developing Science and Health Initiatives (ideSHi) | Hassan Afrad, Sadia Rahman, Manjur Hossain Khan, Firdausi Qadri, Tahmina Shirin |
| EPI_ISL_2180454, EPI_ISL_2180455, EPI_ISL_2180456, EPI_ISL_2180457 | Institute of Epidemiology, Disease Control and Research (IEDCR) | IEDCR-ideSHi-icddr,b | Hassan Afrad, Sadia Rahman, Manjur Hossain Khan, Firdausi Qadri, Tahmina Shirin |
| EPI_ISL_2233084, EPI_ISL_2233085, EPI_ISL_2233086 | Institute for Developing Science and Health Initiatives (ideSHi) | Institute for Developing Science and Health Initiatives (ideSHi) | Hassan Afrad, Sadia Rahman, Manjur Hossain Khan, Firdausi Qadri, Tahmina Shirin |
| EPI_ISL_2233361, EPI_ISL_2233362, EPI_ISL_2233363, EPI_ISL_2233364, EPI_ISL_2233365, EPI_ISL_2233366, EPI_ISL_2233367, EPI_ISL_2233368, EPI_ISL_2233369, EPI_ISL_2233370, EPI_ISL_2233371, EPI_ISL_2233372, EPI_ISL_2233373, EPI_ISL_2233374, EPI_ISL_2233375, EPI_ISL_2233376, EPI_ISL_2233377, EPI_ISL_2233378, EPI_ISL_2233379, EPI_ISL_2233380, EPI_ISL_2233381, EPI_ISL_2235494, EPI_ISL_2235495, EPI_ISL_2235496 | | | |
| see above | Virology Laboratory, International Centre for Diarrhoeal Disease Research, Bangladesh (ICDDR,B) | Virology Laboratory, International Centre for Diarrhoeal Disease Research, Bangladesh (ICDDR,B) | Mohammad Enayet Hossain, Mojnu Miah, Rashedul Hasan, Md. Mahfuzur Rahman, Mohammed Ziaur Rahman, Mustafizur Rahman |
| EPI_ISL_2284885, EPI_ISL_2284886, EPI_ISL_2284887, EPI_ISL_2284888, EPI_ISL_2284889, EPI_ISL_2284890, EPI_ISL_2284891, EPI_ISL_2284892, EPI_ISL_2284893, EPI_ISL_2284894, EPI_ISL_2304461, EPI_ISL_2304462, EPI_ISL_2304463, EPI_ISL_2304465, EPI_ISL_2304466, EPI_ISL_2304467, EPI_ISL_2304468, EPI_ISL_2304469, EPI_ISL_2304470, EPI_ISL_2304471 | | | |
| see above | Institute of Epidemiology, Disease Control and Research (IEDCR) | IEDCR-ideSHi-icddr,b | Hassan Afrad, Sadia Rahman, Manjur Hossain Khan, Firdausi Qadri, Tahmina Shirin |
| EPI_ISL_2348652 | Genome Center | Genome Center | Shovon Lal Sarkar, Md. Shazid Hasan, Ali Ahsan Setu,Tanay Chakrabarty,Prosanto Kumar Das, A. S. M. Rubayet Ul Alam, Ovinu Kibria Islam, Md. Tanvir Islam, Hassan Md. Al-Emran, Iqbal Kabir Jahid, M. Anwar Hossain |
| EPI_ISL_2348658 | Genome Center | Genome Center | Md. Shazid Hasan, A. S. M. Rubayet Ul Alam, Shovon Lal Sarkar, Ali Ahsan Setu, Tanay Chakrabarty,Prosanto Kumar Das, M. Shaminur Rahman, Ovinu Kibria Islam, M. Tanvir Islam, Hassan M. Al-Emran, Iqbal Kabir Jahid, M. Anwar Hossain |
| EPI_ISL_2348660 | Genome Center | Genome Center | A. S. M. Rubayet Ul Alam, Md. Shazid Hasan, Shovon Lal Sarkar, Ali Ahsan Setu, Tanay Chakrabarty,Prosanto Kumar Das, M. Shaminur Rahman, Ovinu Kibria Islam, M. Tanvir Islam, Hassan M. Al-Emran, Iqbal Kabir Jahid, M. Anwar Hossain |
| EPI_ISL_2348661 | Genome Center | Genome Center | M. Tanvir Islam, A. S. M. Rubayet Ul Alam, Md. Shazid Hasan, Shovon Lal Sarkar, Ali Ahsan Setu, Tanay Chakrabarty,Prosanto Kumar Das, M. Shaminur Rahman, Ovinu Kibria Islam, Hassan M. Al-Emran, Iqbal Kabir Jahid, M. Anwar Hossain |
| EPI_ISL_2348662 | Genome Center | Genome Center | Ovinu Kibria Islam, M. Tanvir Islam, A. S. M. Rubayet Ul Alam, Md. Shazid Hasan, Shovon Lal Sarkar, Ali Ahsan Setu, Tanay Chakrabarty,Prosanto Kumar Das, M. Shaminur Rahman, Hassan M. Al-Emran, Iqbal Kabir Jahid, M. Anwar Hossain |
| EPI_ISL_2348663 | Genome Center | Genome Center | Iqbal Kabir Jahid, Md. Shazid Hasan, A. S. M. Rubayet Ul Alam, Shovon Lal Sarkar, Ali Ahsan Setu, Tanay Chakrabarty,Prosanto Kumar Das, M. Shaminur Rahman, Ovinu Kibria Islam, M. Tanvir Islam, Hassan M. Al-Emran, M. Anwar Hossain |
| EPI_ISL_2348664 | Genome Center | Genome Center | Hassan Md. Al-Emran, Shovon Lal Sarkar, Md. Shazid Hasan, Ali Ahsan Setu,Tanay Chakrabarty,Prosanto Kumar Das, A. S. M. Rubayet Ul Alam, Ovinu Kibria Islam, Md. Tanvir Islam, Iqbal Kabir Jahid, M. Anwar Hossain |
| EPI_ISL_2349692 | Genome Center | Genome Center | Ovinu Kibria Islam, Hassan Md. Al-Emran, Shovon Lal Sarkar, Md. Shazid Hasan, Ali Ahsan Setu,Tanay Chakrabarty,Prosanto Kumar Das, A. S. M. Rubayet Ul Alam, Md. Tanvir Islam, Iqbal Kabir Jahid, M. Anwar Hossain |
| EPI_ISL_2349879 | Genome Center | Genome Center | Md. Tanvir Islam, Hassan Md. Al-Emran, Shovon Lal Sarkar, Md. Shazid Hasan, Ali Ahsan Setu,Tanay Chakrabarty,Prosanto Kumar Das, A. S. M. Rubayet Ul Alam, Ovinu Kibria Islam, Iqbal Kabir Jahid, M. Anwar Hossain |
| EPI_ISL_2349880 | Genome Center | Genome Center | Hassan Md. Al-Emran, Shovon Lal Sarkar, Md. Shazid Hasan, Ali Ahsan Setu,Tanay Chakrabarty,Prosanto Kumar Das, A. S. M. Rubayet Ul Alam, Ovinu Kibria Islam, Md. Tanvir Islam, Iqbal Kabir Jahid, M. Anwar Hossain |
| EPI_ISL_2350142 | Genome Center | Genome Center | A. S. M. Rubayet Ul Alam, Md. Shazid Hasan, Shovon Lal Sarkar, Ali Ahsan Setu, Tanay Chakrabarty,Prosanto Kumar Das, M. Shaminur Rahman, Ovinu Kibria Islam, M. Tanvir Islam, Hassan M. Al-Emran, Iqbal Kabir Jahid, M. Anwar Hossain |
| EPI_ISL_2350167 | Genome Center | Genome Center | Md. Shazid Hasan, Shovon Lal Sarkar, Ali Ahsan Setu, A. S. M. Rubayet Ul Alam, Tanay Chakrabarty,Prosanto Kumar Das, M. Shaminur Rahman, Ovinu Kibria Islam, M. Tanvir Islam, Hassan M. Al-Emran, Iqbal Kabir Jahid, M. Anwar Hossain |
| EPI_ISL_2361892, EPI_ISL_2361893, EPI_ISL_2361894, EPI_ISL_2361895, EPI_ISL_2361896, EPI_ISL_2361897, EPI_ISL_2361903 | Virology Laboratory, International Centre for Diarrhoeal Disease Research, Bangladesh (ICDDR,B) | Virology Laboratory, International Centre for Diarrhoeal Disease Research, Bangladesh (ICDDR,B) | Mohammad Enayet Hossain, Mojnu Miah, Rashedul Hasan, Md. Mahfuzur Rahman, Mohammed Ziaur Rahman, Mustafizur Rahman |
| EPI_ISL_2399440, EPI_ISL_2399441, EPI_ISL_2399442, EPI_ISL_2399443, EPI_ISL_2399444, EPI_ISL_2399445, EPI_ISL_2399446, EPI_ISL_2399447, EPI_ISL_2399448, EPI_ISL_2399449, EPI_ISL_2399450, EPI_ISL_2399451, EPI_ISL_2399452, EPI_ISL_2399453, EPI_ISL_2399454, EPI_ISL_2399455, EPI_ISL_2399456 | | | |
| see above | Institute of Epidemiology, Disease Control and Research (IEDCR) | IEDCR-ideSHi-icddr,b | Hassan Afrad, Sadia Rahman, Manjur Hossain Khan, Firdausi Qadri, Tahmina Shirin |
